# Upregulated miR-200c may increase the risk of obese individuals to severe COVID-19

**DOI:** 10.1101/2021.03.29.21254517

**Authors:** Jayanthi Bellae Papannarao, Daryl O Schwenke, Patrick Manning, Rajesh Katare

## Abstract

Obesity is a risk factor for coronavirus disease 2019 (COVID-19) infection, the prevalence of obese individuals admitted with COVID-19 ranging between 30 and 60%. Herein we determined whether early changes in microRNAs (miRNAs) could be the underlying molecular mechanism increasing the risk of obese individuals to COVID-19 infection. Quantitative real-time PCR analysis of plasma samples for circulating miRNAs showed a significant upregulation of miR-200c and a small increase in miR-let-7b obese individuals. This was associated with significant downregulation of angiotensin-converting enzyme 2 (ACE2). Both the miRNAs are the direct targets of ACE2, the specific functional receptor for severe acute respiratory syndrome coronavirus 2. Correlation analysis confirmed a significant negative correlation between ACE2 and both the miRNAs. Recent studies showed that despite being the functional receptor, inhibition/downregulation of ACE2 did not reduce the severity of COVID-19 infection. In contrast, increased angiotensin II following inhibition of ACE2 may increase the severity of the disease. Taken together, our novel results identify that upregulation of miR-200c may increase the susceptibility of obese individuals to COVID-19. Considering miRNA are the earliest molecular regulators, circulating miR-200c could be a potential biomarker in the early identification of those at the risk of severe COVID-19.

---

Obesity is considered by many as an epidemic of the 21^st^ century, with recent analysis suggesting that obesity prevalence could reach 20% of the world’s population by 2025^1^. Obesity is associated with the development of type 2 diabetes mellitus, cardiovascular diseases including stroke and hypertension, certain types of cancers, and several other pathological conditions^2,3^. Severe acute respiratory syndrome coronavirus 2 (SARS-CoV-2) causing coronavirus disease 2019 (COVID-19) pandemic has a devastating and chronic effect on the health of an individual, killing more than 2.6 million people (to date) worldwide. Dissemination of a vaccine and immunization program against COVID-19 is likely to reduce the intensity and mortality rate in most cohorts of patients without any other risk factors. However, people who fall within a high-risk category, such as obesity, still remain susceptible to ongoing complications associated with COVID-19. Obesity is considered a risk factor for COVID-19 infection, with studies demonstrating the prevalence of obese individuals admitted with COVID-19 ranging between 30 and 60%^4^. Once infected, obese individuals develop a serious illness associated with a higher mortality rate^4^. Excessive adiposity is thought to be causally linked to this high mortality with COVID-19 infection^5^ because increased fat disrupts normal metabolism, activates multiple inflammatory cytokines and reduces anti-inflammatory agents such as adiponectin, all of which normally protect the lung against infection. Importantly, the exact molecular mechanism(s) still remain elusive, yet are essential for furthering our understanding and establishing preventative strategies against the insult of COVID-19.

MicroRNAs (miRNAs) are small non-coding RNA molecules that constitute 1-5% of the human genome and regulate at least 30% of all protein-coding genes^6^. Accumulating evidence from the past decade has identified miRNAs as critical elements in several physiological and pathological processes^6^. Further, miRNAs are secreted into the circulation, where they remain stable, hence serving as a valuable diagnostic and prognostic biomarker of diseases^6-8^. Our recent study identified dysregulation of circulating miRNAs associated with diabetes, insulin signaling, cell survival, and cardiovascular diseases in obese individuals^9^. Interestingly, the majority of the miRNAs were normalized following acute weight loss in these individuals, demonstrating the role of miRNAs as biomarker^9^. In support of this, other studies have demonstrated the diagnostic potential of miRNAs in cardiovascular diseases, diabetes, and cancer^6-8^.

In this study, we aimed to determine whether obesity-induced dysregulation of miRNAs could be the underlying mechanism for increasing the risk for developing COVID-19 infection in obese individuals. In particular, we tested two miRNAs, miR-200c and miR-let-7b, both of which directly target angiotensin-converting enzyme 2 (ACE2), the specific functional receptor for SARS-CoV-2^10^.

Plasma samples were collected from obese individuals who were participants in the ongoing Prevention Of WEight Regain (POWER) study (Clinical Trials Registry number: ACTRN12612000069853), which was designed to assess the effects of cabergoline on the prevention of weight regain in obese individuals was used for the current study. All participants in the POWER study provided plasma samples at baseline. Plasma samples from age-matched lean individuals were used as control (*detailed below in Methods*).

Baseline clinical data showed significantly higher BMI and blood pressure in obese individuals (**Table 1**). Quantitative real-time PCR analysis of the total RNA isolated from plasma showed a significant upregulation of circulating miR-200c in obese individuals compared to the lean individuals (P<0.0001 vs. lean, **Figure 1A**). While miR-let-7b expression was higher in obese individuals, it was not statistically (P=0.312, **Figure 1B**). ELISA revealed a significant decrease in the level of secreted ACE2, the direct target of miR-200c and miR-let-7b in obese individuals (P=<0.0001, **Figure 1C**).

**Table 1:**
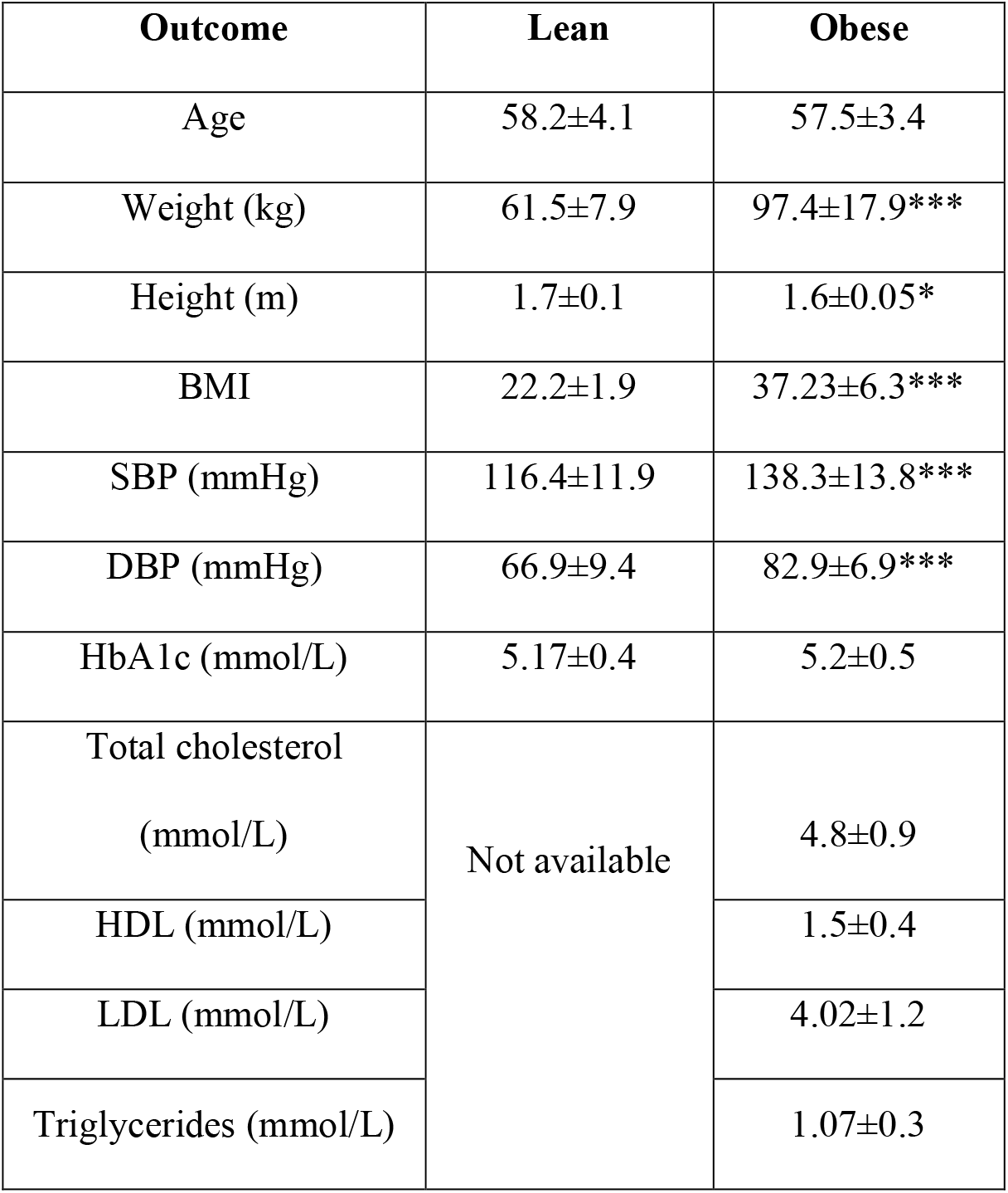
Anthropometric and metabolic parameters in study participants. BMI – body mass index; SBP – systolic blood pressure; DBP – diastolic blood pressure; HbA1c – glycated haemoglobin; HDL – high density lipoprotein; LDL – low density lipoprotein. *P<0.05 and ***P<0.0001 vs. lean group.

| **Outcome** | **Lean** | **Obese** |
| --- | --- | --- |
| Age | 58.2±4.1 | 57.5±3.4 |
| Weight (kg) | 61.5±7.9 | 97.4±17.9*** |
| Height (m) | 1.7±0.1 | 1.6±0.05* |
| BMI | 22.2±1.9 | 37.23±6.3*** |
| SBP (mmHg) | 116.4±11.9 | 138.3±13.8*** |
| DBP (mmHg) | 66.9±9.4 | 82.9±6.9*** |
| HbA1c (mmol/L) | 5.17±0.4 | 5.2±0.5 |
| Total cholesterol<br>(mmol/L) | Not available | 4.8±0.9 |
| HDL (mmol/L) |  | 1.5±0.4 |
| LDL (mmol/L) |  | 4.02±1.2 |
| Triglycerides (mmol/L) |  | 1.07±0.3 |

**Figure 1.**
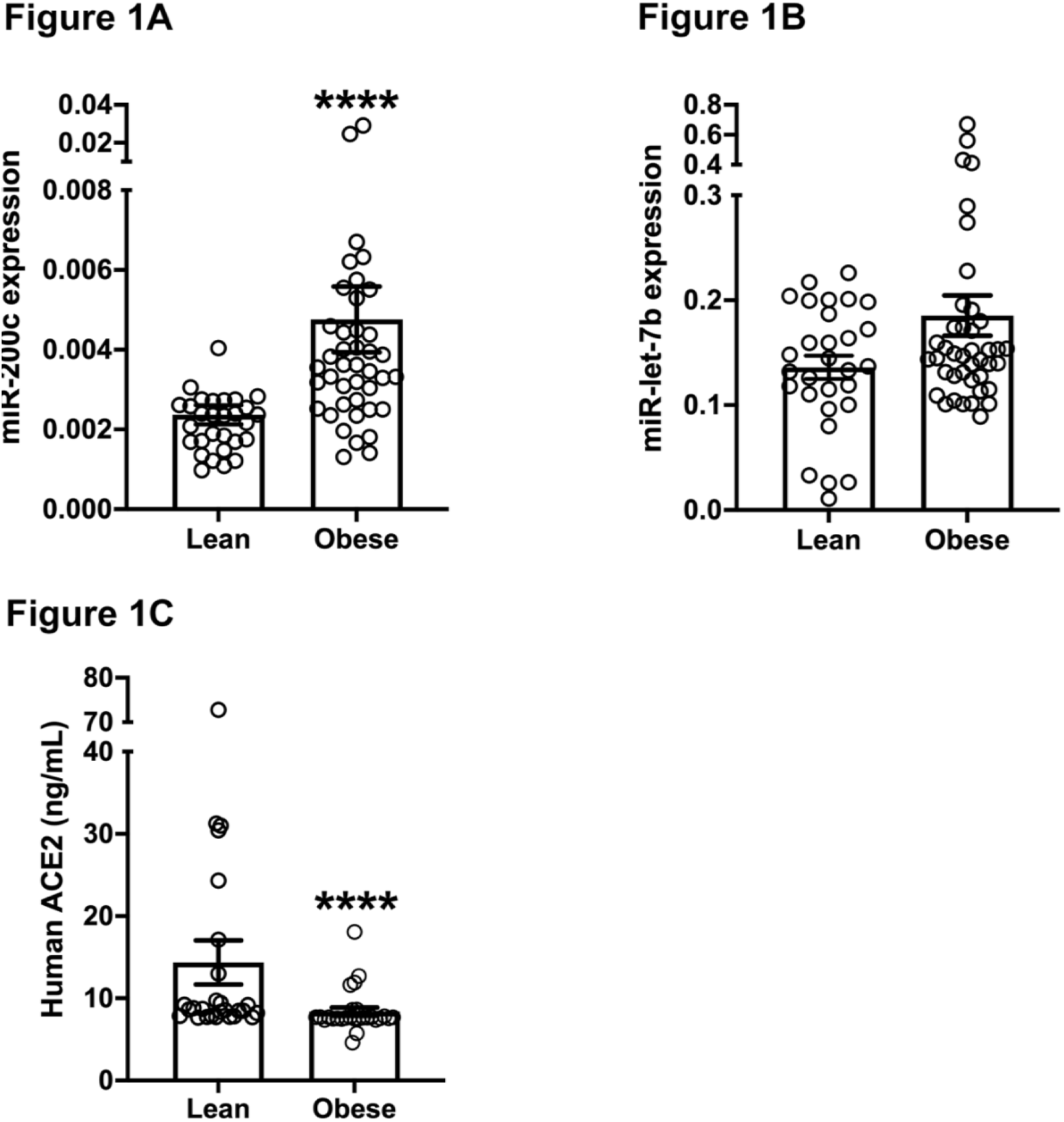
**A&B** – Quantitative scatter plot bar graphs showing the RT-PCR analysis of miR-200c (**A**) and let-7b (**B**) in the study participants. All the experiments were repeated at least 2 independent times. **C**. Quantitative scatter plot bar graph showing human ACE2 levels in study participants measured by ELISA. ELISA was performed in triplicates. The data are presented as mean±SEM. N= 30 in lean and 31 in obese individuals. ****P<0.0001 vs. lean participants.

Next, to determine the correlation between miR-200c upregulation and reduced levels of ACE2, we used the Spearman correlation coefficient, which showed a significant negative correlation between miR-200c and ACE2 irrespective of the group (r = -0.3054, P=0.0247, **Figure 2B**). Interestingly, although miR-let-7b expression was not significantly different in obese subjects, correlation analysis showed a significant negative correlation between miR-let-7b and ACE2 (r = -0.3061, P=0.0258, **Figure 2C**).

**Figure 2.**
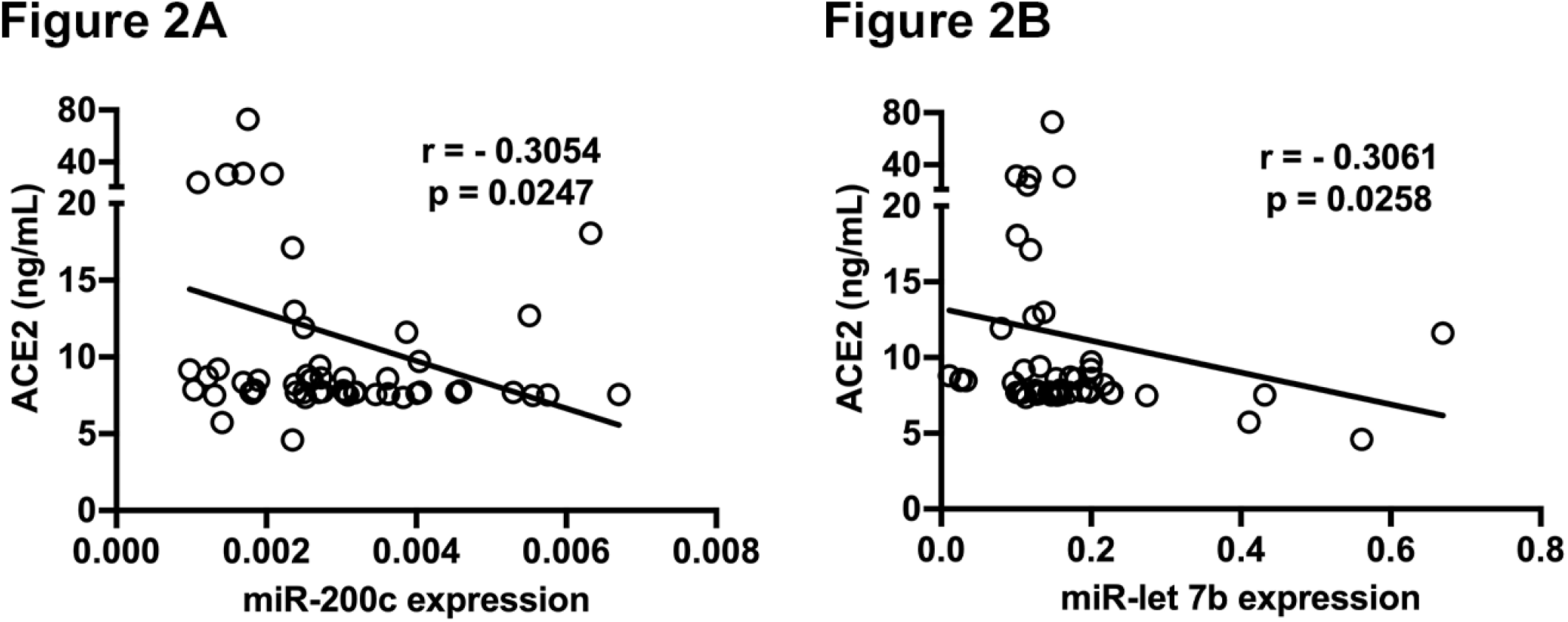
Line graphs with scatter plots of Spearman correlation analysis showing significant negative correlation between ACE2 and miR-200c (**A**) and let-7b (**B**).

A crucial finding from our study is that upregulation of miR-200c associated with obesity could be a molecular mechanism increasing the risk in individuals. miR-200c is located in the intergenic region of chromosome 12, along with miR-141, and is highly expressed in epithelial cells, including type 2 alveolar epithelial cells^11^. In the fetal lung, upregulation of miR-200c contributed to the differentiation of type II alveolar epithelial cells^11^. On the other hand, a higher miR-200c expression was associated with shorter survival in patients with non-small cell lung cancer^12^. Further, miR-200c directly targets ACE2, and our results confirmed a strong negative correlation between the miR-200c and ACE2. Interestingly, miR-let-7b, another known modulator of ACE2, did not reach significance in obese individuals, which is likely due to increased variability in miR-let-7b expression among study participants. However, it showed a significant negative correlation to ACE2.

Being the main receptor for SARS-CoV-2, ACE2 is considered a promising therapeutic target for the treatment of COVID-19^13^. Therefore, it is intriguing to consider that increased miR-200c may be beneficial in the obese individual by directly targeting ACE2, which is supported by our results. However, obesity is strongly and independently associated with adverse outcomes of COVID-19^5^. ACE2 is expressed in nearly all human organs in varying degrees and plays a crucial role in cellular homeostasis. In the lungs, like miR-200c, ACE2 is mainly expressed on type II alveolar epithelial cells. It is an important component of the renin-angiotensin system (RAS) signaling throughout the body and exhibits anti-inflammatory, anti-remodeling, and anti-proliferative properties through reduction of angiotensin II levels. It is important to note that COVID-19 patients using angiotensin-converting enzyme inhibitors or angiotensin receptor blockers, which upregulate ACE2 level, do not show higher mortality rates or increased disease severity^14^. Therefore, having a low level of ACE2 is unlikely to be beneficial. On the other hand, when individuals with the low level of ACE2, such as the obese individuals in our study, is affected with COVID-19, binding of SARS-CoV-2 to ACE2 is likely to lead to reduced ACE2 cell surface expression, resulting in critically low ACE2 levels and exaggerated angiotensin II signaling, leading to severe form of the disease^15^.

While it can be argued that measurement of ACE2 level is sufficient to identify those at the risk of developing severe disease, dysfunction in miRNAs always precede changes in protein and cellular structure/function and, thus, are the earliest molecular regulators. We, along with others, have demonstrated the changes in miRNA well before the changes are identified at the target protein level^6-8^. Therefore, the early detection of increased miR-200c expression in obese subjects, from a simple blood test, could significantly aid in mitigating the risk of contracting COVID-19. This study focused on obese individuals, however, it will be interesting to determine the level of miR-200c in individuals affected with COVID-19 to establish its role as a biomarker to identify those at risk of contracting COVID-19.

## Methods

### Participants and sample collection

The Health and Disability Ethics Committee of New Zealand approved the use of samples for this study. Participants were identified from the ongoing Prevention Of WEight Regain (POWER) study (Clinical Trials Registry number: ACTRN12612000069853), which was designed to assess the effects of cabergoline on the prevention of weight regain in obese individuals. All participants in the POWER study provided plasma samples at baseline^9,16^. Informed consent was obtained from each patient for the collection and use of samples in this study, and the study protocol conforms to the ethical guidelines of the 1975 Declaration of Helsinki as reflected in a priori approval by the New Zealand Health and Disability Ethics Committee. Forty-one female participants for the present study were randomly selected from the 221 female individuals recruited in this study. Men were excluded as there were very few recruited into the POWER study. Plasma samples were also collected from 29 age-(±1.5 years) matched female lean individuals recruited through public advertising. Exclusion criteria for both lean and obese participants were pregnancy, diabetes, malignancy, cardiac disease, other vascular diseases, uncontrolled hypertension, psychiatric disease, or any other significant medical condition.

### RNA extraction and quantitative real-time PCR analysis

Total RNA was extracted from plasma using QIAgen miRNAeasy mini kit following the manufacturer’s instruction and described in our earlier studies^6-8^. Ten nanograms of total RNA were reverse transcribed using miR-200c and let 7b and internal controls miR-16 and miR-24 specific stem-loop structure and reverse transcription primers (all from Thermofisher Scientific). The resulting cDNA was then amplified using specific Taqman hybridization probes to quantify the expression of tested miRNAs (BioRad Laboratories). miRNA expression was normalized to internal controls and presented as DCT (2^−ΔCt^) expression. All the experiments were repeated at least 2 independent times, and the data from each independent repeat were averaged.

### ELISA for ACE2

Plasma (50µL) from obese and lean individuals was used to measure the level of ACE2 using the commercial kit (Human ACE-2 DuoSet ELISA, R&D Systems) following the manufacturer’s instructions.

### Statistical analysis

All statistical analyses were performed using GraphPad Prism (version 8). Shapiro–Wilk test was used to test the normality. A non-parametric Mann–Whitney U test was used to analyze the quantitative real-time PCR analysis and ELISA results. Data are expressed as the mean ± standard error of the mean (SEM). Spearman correlation analysis was used to determine the correlation between ACE2 and miRNAs. A p-value < 0.05 was considered statistically significant.

## Data Availability

All the data are available with the corresponding author on request.

## Acknowledgments

This study was supported by funding from Lottery Health Research Funding (R-LHR-2020-128280), the University of Otago Research Funding and the Department of Physiology funding.

## Author contributions

R.K., J.B.P., and D.O.S., designed the research; P.M provided access to the study samples; J.B.P. performed the research; R.K. and J.B.P. analyzed the data; R.K, D.O.S., and P.M. wrote the paper.

